# A Systematic review of ‘Fair’ AI model development for image classification and prediction

**DOI:** 10.1101/2022.05.18.22275254

**Authors:** Ramon Correa, Mahtab Shaan, Hari Trivedi, Bhavik Patel, Leo Anthony G. Celi, Judy W. Gichoya, Imon Banerjee

## Abstract

**Background:** Artificial Intelligence (AI) models have demonstrated expert-level performance in image-based recognition and diagnostic tasks, resulting in increased adoption and FDA approvals for clinical applications. The new challenge in AI is to understand the limitations of models to reduce potential harm. Particularly, unknown disparities based on demographic factors could encrypt currently existing inequalities worsening patient care for some groups.

**Method:** Following PRISMA guidelines, we present a systematic review of ‘fair’ deep learning modeling techniques for natural and medical image applications which were published between year 2011 to 2021. Our search used Covidence review management software and incorporates articles from PubMed, IEEE, and ACM search engines and three reviewers independently review the manuscripts.

**Results:** Inter-rater agreement was 0.89 and conflicts were resolved by obtaining consensus between three reviewers. Our search initially retrieved 692 studies but after careful screening, our review included 22 manuscripts that carried four prevailing themes; ‘fair’ training dataset generation (4/22), representation learning (10/22), model disparity across institutions (5/22) and model fairness with respect to patient demographics (3/22). However, we observe that often discussion regarding fairness are also limited to analyzing existing bias without further establishing methodologies to overcome model disparities. Particularly for medical imaging, most papers lack the use of a standardized set of metrics to measure fairness/bias in algorithms.

**Discussion:** We benchmark the current literature regarding fairness in AI-based image analysis and highlighted the existing challenges. Based on the current research trends, exploration of adversarial learning for demographic/camera/institution agnostic models is an important direction to minimize disparity gaps for imaging. Privacy preserving approaches also present encouraging performance for both natural and medical image domain.

## 1 Introduction

The development of Artificial intelligence (AI) models, particularly deep learning, in medical imaging has achieved remarkable performance such that models are able to match expert-level accuracy. For example, performance at par with specialists has been shown in a variety of clinical applications, from diagnosing common thoracic pathologies on chest radiographs Rajpurkar et al. (n.d.) to detecting diabetic retinopathy on retinal fundoscopic images Ting et al. (n.d.). Interestingly, some recent studies have claimed that AI has surpassed expert-level performance. For example, Becker et al. (n.d.) showed that AI had a higher sensitivity for detecting abnormalities on screening mammograms when compared to a radiologist, particularly for subtle lesions. Lee et al. (2020) showed that analysis of brain MRI using AI has greater sensitivity in identifying parenchymal changes reflective of early ischaemic stroke within a narrower time window from onset of symptoms with greater sensitivity than a human reader.

However, in parallel with this remarkable success, recent works have raised concerns towards the risk of unintended bias in AI systems affecting individuals unfairly based on race, gender, and other clinical characteristics Seyyed-Kalantari et al. (2021); Parikh et al. (n.d.); Whittaker et al. (n.d.). AI bias is defined as models with outputs providing outcomes that negatively affects one sub-group of the study population more than others. Examples include differing allocation of healthcare resources based on patient demographics Obermeyer et al. (2019); Benjamin (2019), bias in language models developed on clinical notes Zhang et al. (2020), and melanoma detection models developed primarily on images of light-colored skin Adamson and Smith (2018). Beyond healthcare applications, similar biases have been observed for tasks face detection where models fail to correctly identify individuals of minority groups Buolamwini and Gebru (n.d.), and achieved lower performance for cyberbully detection for unprivileged group.

A core challenge for analyzing AI bias is that the reasons resulting in unfairness of AI models are not mutually exclusive and can often exacerbate one another. Recently, Banerjee et al. (2021) showed that AI models trained for diagnosis can learn unintended racial information from different imaging modalities. Thus, AI models may use learned demographic information for detecting a diagnosis even when such attribute is not associated with the diagnosis. There are examples of race-ethnicity and gender influencing clinician decision-making Wallis et al. (2022), and given that AI is trained on real-world data, it is not far-fetch to assume that computers will learn to do the same.

Medical imaging datasets used to train and validate algorithms tend to originate from single health systems where the patient demographics may only be representative of a segment of the population creating data deserts without representation in AI training data Kaushal et al. (2020). Moreover, clinical practice patterns, which influence the relationship between patient and disease features with diagnosis and clinical outcomes, vary across health systems and over time. Local practice pattern changes and other extraneous factors, such as adoption of new clinical protocols, are not usually reflected in the EMR. This often brings into question how reliable models generalize to populations across space and time, how these models might fail, and whether they will entrench or even scale health disparities. Previous works have only sought to study the generalization of model performance across institutions without consideration of demographic shifts across institutions or over time Davis et al. (2020). There is a critical need to evaluate how individual, institutional and societal biases can be mitigated or exacerbated by adopting AI models.

To our knowledge, no systematic review has aggregated information regarding imaging-based AI models contributing to disparities and methods of systematically reducing model bias. The review will identify existing best practices in developing AI models for both natural and clinical images as well as gaps in the evaluation of and mitigation of the impact of algorithms on outcome inequities. In addition, we also highlight the challenges on how the best practices will be implemented and how the gaps will be addressed.

### 1.1 Terminologies

We briefly define the terms and their acronyms that have been referenced throughout the paper as well as in the ‘fair’ AI field.

- *‘Fair’ AI model*. Ideally, a ‘Fair’ AI model is defined as a model without any disparate treatment and disparate impact to certain unpledged groups based on protected attributes like gender, race, religion, color, age, and more.
- *Model Bias*. In statistics, bias is defined as an error generated from erroneous assumptions used in the learning algorithms. There are two types of bias that can affect AI models. One is ‘algorithmic AI bias’ where algorithms are trained using biased data while ‘societal AI bias’ is caused by the assumptions and norms imposed by the society. For example, AIs trained on news articles show a bias against women. Societal bias often significantly influences algorithmic AI bias.
- *Model loss and optimization*. In brief, a machine learning model learns to map a set of inputs to a set of outputs from training data using the best weights. Learning which weights are the best for learning is achieved through search optimization by navigating a set of possible weights towards the most optimal task performance.
- *Performance measures, AUC, mAP, MAE*. Model evaluation aims to estimate the generalization accuracy of a model on unseen test data. Receiver operating characteristic curve (ROC) plots true positive rate vs. false positive rate at different classification thresholds. AUC measures the entire two-dimensional area underneath the ROC curve and is commonly measured for binary classification problems. The mean Average Precision or mAP score is calculated by taking the average precision over all classes. Mean absolute error (MAE) is a measure of errors between the true and model predicted values, and is primarily used for regression task.
- *Model Disparity*. Disparity can be defined as difference in model performance across different population subgroups. Inadequate training data may lead to suboptimal AI models with high performance for majority of training data but inherent bias for minority groups, which may have profound negative impacts on health care.
- *Disparity measures*. Common metrics to measure the model disparities include difference in true positive rate (TPR disparity) and false negative rate (FNR disparity) across different subgroups against the reference group (majority class).

## 2 Method

We conducted a systematic review of ‘fair’ AI model development in imaging following the PRISMA guidelines Moher et al. (2009). A search for natural images included images from landscape, person, as well as aerial, cartoon, and LiDAR images. Medical imaging search included all studies of images related to various body-parts acquired for diagnostic and treatment purposes, e.g: X-ray, H&E slides, magnetic resonance images. The following subsections respectively describes our search criteria, results, and summary.

### 2.1 Search strategy

Our search was obtained by querying PubMed, IEEE’s, and ACM’s engines and reviewed by the team in Covidence. The results are aggregated together and screened as specified in Figure 1. The search query was “(disparity OR bias OR fairness) AND (classification OR prediction) AND (image) AND (deep learning[MeSH Terms])” present within the abstract of the article. The queries were limited to the last ten years of publication (Jan, 2011 July, 2021) and the systems developed to support image classification and prediction tasks. Models for image segmentation and object detection task were excluded due to distinct methodological challenges for bias removal.

**Figure 1.**
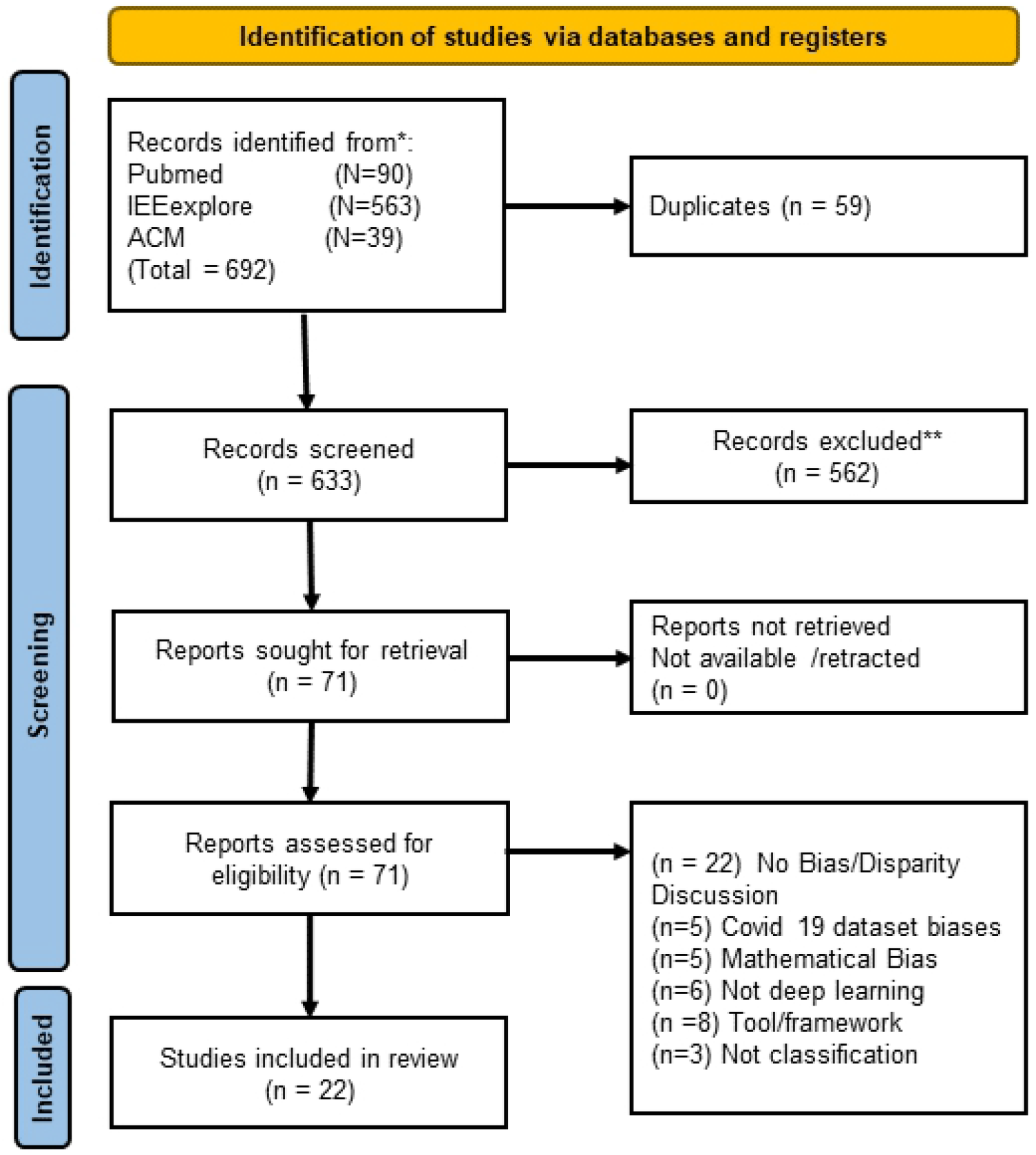
PRISM flowchart for literature screening. n represents number of unique papers.

### 2.2 Study selection

Our study screening process involved *three independent reviewers* who screened a total of 692 studies retrieved by the initial search in Covidence. Interreader agreement was assessed using Fleiss’ kappa score. Papers were excluded if they met one of the following criteria i) no peer review (e.g. published in arXiv, medRxiv, institutional databases); ii) the main topic was dataset curation and no model development and training was proposed; (ii) not a deep learning model. 71 papers were assessed for eligibility and, finally, included the papers which either discuss the disproportionate performance of models and/or propose novel techniques for mitigating model bias.

## 3 Results

Figure 1 presents the PRISM selection flowchart. Inter-rater agreement was 0.89 and conflicts were resolved by obtaining consensus between three reviewers. Based on the consensus of all three reviewers, 22 papers satisfied our inclusion criteria and were selected for final extraction and review. A summary of the selected literature is presented in 3 tables - studies applied to natural images: Table 1, 2, and medical images: Table 3. Figure 2 shows the diagrammatic representation of the study grouping strategy for this review. We adopted five benchmarking criteria to analyze the papers - (1) *Dataset origin*: training and evaluation data, (2) *Modality* : targeted imaging modality, (3) *Method for de-biasing* : strategy for mitigating bias, (4) *Targeted task* : actual prediction or classification objective, and (5) *Performance*: final model performance reported (after de-biasing if novel mitigation strategy proposed). We present a summary of the papers in terms of model fairness in the *Observation* column.

**Table 1:**
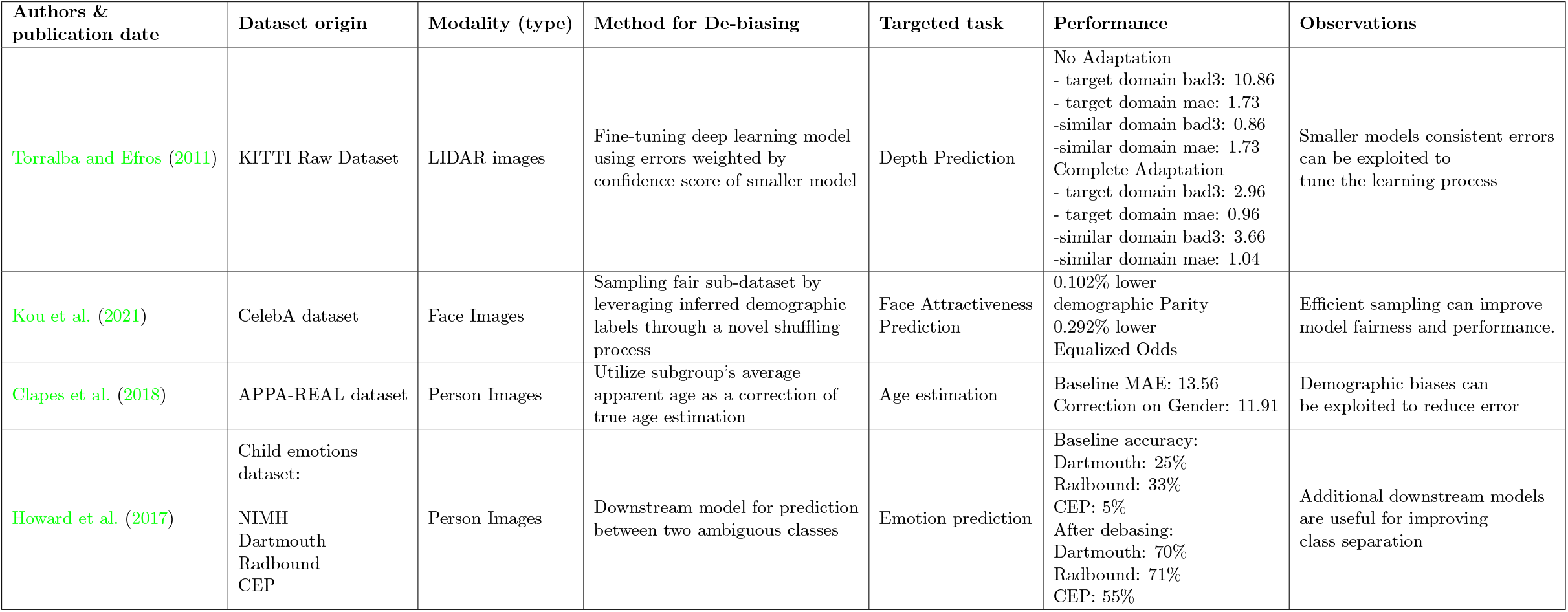
Benchmarking articles related to proposing new dataset sub-sampling techniques for reducing bias.

**Table 2:** Benchmarking articles related to proposing new techniques for ‘fair’ representation learning from generic images.

| Authors & publication date | Dataset origin | Modality (type) | Method for De-biasing | Targeted task | Performance | Observations |
| --- | --- | --- | --- | --- | --- | --- |
| Morales et al. (2020) | Dive Face dataset | Facial Images | Adversarial regularizer. | Face identification | Adverse task:<br>Gender: before 97.7, after: 58.8<br>Ethnicity: before 98.8%, after 55.1% | Privacy preserving representations learning for bias reduction. |
| Alsulaimawi (2020) | CelebA | Facial images | Feature protection using adversarial training | Smile detection | Target: AUC 0.97<br>Advers(*): AUC: .51 | Autoencoders can remove irrelevant features allow training a fair classifier. |
| Quadrianto et al. (2019) | CelebA, DiF | Facial images | Unconstrained mapping using residual decomposition | Face attractiveness prediction & Age prediction | CelebA<br>Baseline acc:80.6<br>Proposed: acc:79.4 | Removal of biased features can reduce biased outcomes. |
| Zhang et al. (2021) | Market1501, DukeMTMC-reID, MSMT17 | Person Images | Unsupervised multi-scale self-training for view-invariant representation learning | Person Re-identification | Dataset increase:<br>avg 3.16% mAP, 5.16% in rank accuracy | confounding features can be separable during adversarial debiasing. |
| Yu et al. (2018) | ViPeR ,CUHK0, CUHK0, SYSU, Market-1501, ExMarket | Person images | Asymmetric metric learning | Person identification | ExMarket:<br>-DIC model MAP 42.18 (21.19)<br>-DECAMEL model 62.98 (33.28) | Effect of task irrelevant data clusters can be reduced using distance-based loss. |
| Jiang et al. (2019) | Private Dataset | Person Images | Attribute aware loss | Face identification | Accuracy<br>RGB: 87.50<br>RGB+Depth: 92.84<br>RGB+Depth+attribute: 96.5 | Task performance can improve by including demographic attributes in training. |
| Yan et al. (2019) | WHU-RS,SIRI-WHU,jinmen | Aerial images | image to subcategory distance function | classification of land cover | Baseline (res):<br>92.99, 88.73, 87.01<br>Proposed :<br>96.69, 94.50, 91.74 | Additional loss can enforce similarity across domains. |
| Adeli et al. (2021) | Private Brain MRI dataset<br>Gender Shades Pilot Parliaments Benchmark | Brain MRI<br>Person Images | Domain adaptation via statistical independence | Prediction of HIV status<br>Gender prediction | MRI (AUC,corr) : (80.9,.05)<br>Gender:<br>BR-NET: 99.4,.12 | Learn better representations via reduced correlation between features & confounding task |
| Tonioni et al. (2019) | KITTI Raw Dataset | LIDAR images | Domain adaptation via model fine-tuning | Depth Prediction | Baseline<br>MAE 1.73<br>Complete Adaptation<br>MAE:0.96 | Related task can provide additional error estimation during the adaptation process |
| Li et al. (2017) | PACS (Photo, Art , Cartoon, Sketch), VLCS dataset | Photo, Art, Cartoon, Sketch | Domain generalization based on low-rank parameterized CNNs | Non photo-realistic image classification | VLCS proposed MAP: 72.11%<br>Baseline MAP 72.02%<br>PACS proposed MAP: 69.21%<br>Baseline: 67.37% | Learning a subset of parameters to be domain specific improves generalization |

**Table 3:** Benchmarking articles related to proposing new debiasing techniques for medical imaging.

| Authors & publication date | Dataset origin | Modality | Method for de-biasing | Targeted task | Performance | Observations |
| --- | --- | --- | --- | --- | --- | --- |
| Institutional Bias |  |  |  |  |  |  |
| Hägele et al. (n.d.) | TCGA SKCM<br>TCGA-BRCA<br>TCGA-LUAD<br>-open source | Digital Pathology | Remove biasing data samples | Cancer cell detection | Improve AUC by 0.05 with de-biasing | Model interpretation technique to discover hidden bias. |
| D. Das et al. (n.d.) | Montgomery County Hospital, USA & Shenzhen Hospital China<br>- private | Chest X-ray | Dataset consolidation | Predicting chest X-ray findings | AUC:0.84 | Performance improved across sites by balancing prevalence of diseases. |
| Zech et al. (n.d.) | Mount Sinai Hospital and Indiana University, USA<br>- Private | Chest X-ray | Dataset balancing with equal site incidence | Diagnosis of abnormalities in chest X-ray | AUC:0.815 | Balancing proportions of site incidence achieved improved performance |
| Dinsdale et al. (n.d.) | UK BioBank<br>OASIS dataset (healthy only)<br>Whitehall II Study (healthy Only)<br>- open-source | Brain MRI | Confusion Loss | Age Estimation | Dataset MAE:(Original,Unlearning)<br>Biobank: 3.24, 3.38<br>OASIS: 4.19,3.90<br>Whitehall: 2.89 ,2.56<br>Scanner Accuracy: 96,34 % | Separation of correlated task and confounder features through selection of data subsets. |
| Demographic Bias |  |  |  |  |  |  |
| Suriyakumar et al. (n.d.) | NIH-Chest X-ray<br>- open-source | Chest X-ray | Privacy preserving training | Predicting chest x-ray findings | AUC:0.84 | Differential privacy techniques suffer performance decrease despite theoretical guarantees. |
| Larrazabal et al. (n.d.) | NIH Chest X-ray<br>- open-source | Chest X-ray | Balancing proportions for demographic factors | Diagnosis of abnormalities in chest X-ray | AUC: 0.81 | Training on equal proportion of gender improves model performance. |
| Seyyed-Kalantari et al. (n.d.) | NIH Chest X-ray,<br>MIMIC chexpert<br>- open-source | Chest x-ray | Observational,<br>no debiasing | Diagnosis of abnormalities in chest X-ray | Sex<br>TPR GAP: .045<br>Age<br>TPR GAPR: .215<br>Race TPR GAP: .226 | Positive correlation exist between diseases incidence and demographic disparity but not statistically significant |
| Guenther et al. (n.d.) | EU BIOBank<br>- open-source | Retinal fundus | Observational,<br>no debiasing | Age related macular degeneration classification | %FP light eyes: 1.2 ,<br>%FP in dark eyes: 3. | Statistical analysis of misclassification; identified darker eye image to be misclassified more often. |

**Figure 2.**
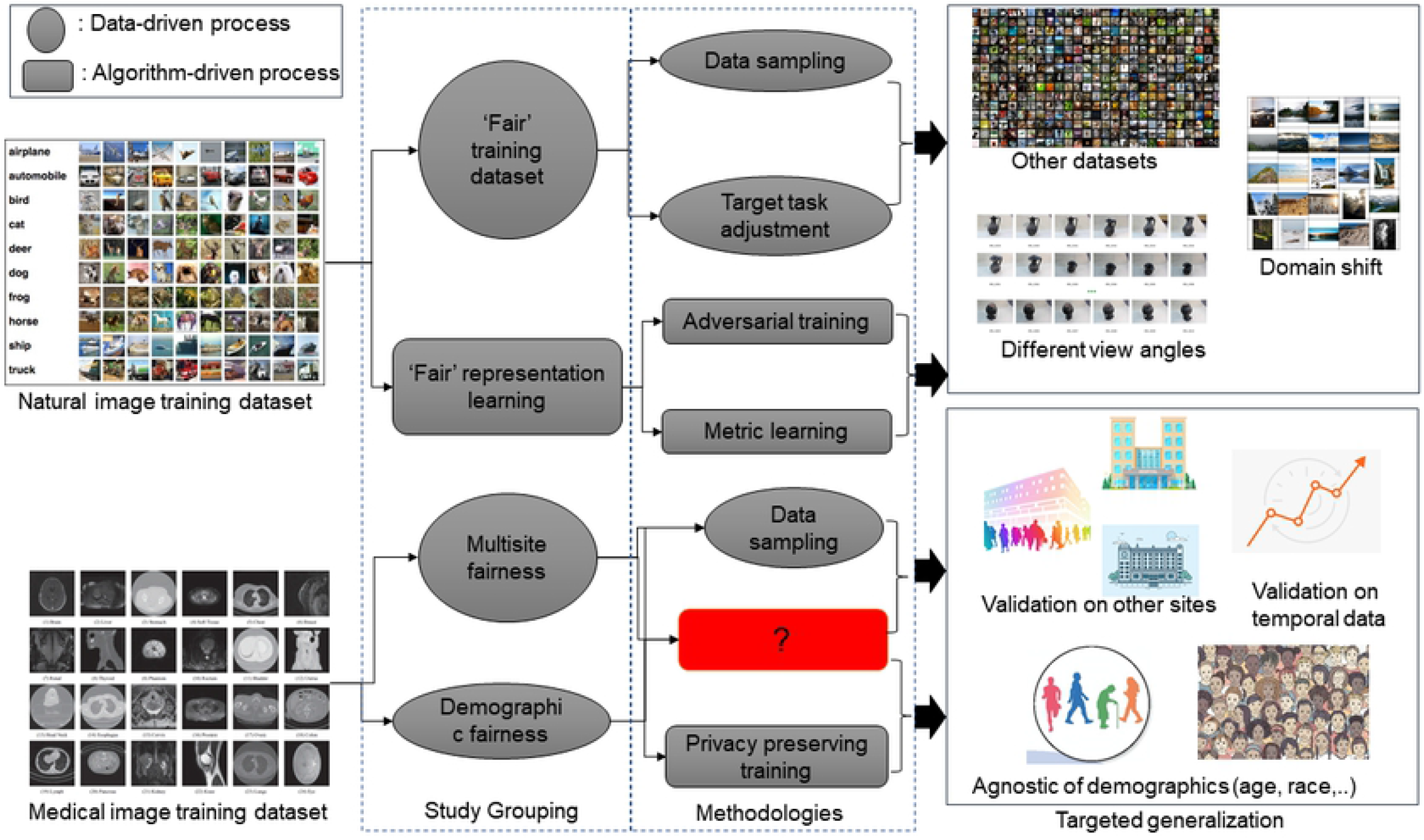
Pictorial representation of study grouping strategy: natural images and medical images.

### 3.1 Natural imaging study grouping

We grouped the debiasing of deep learning models proposed for natural imaging into two core method themes - (1) *data sampling to reduce implicit training data bias* and (2) *fair representation learning for better understanding of data*.

#### 3.1.1. ‘Fair’ training dataset

Often bias introduce in the model due to certain elements are more heavily weighted and/or represented than others within the training samples which resulted skewed outcomes, low accuracy levels. We discuss the articles that focused on developing intelligent data sampling or target task adjustment for reducing the inherent bias present in the training data.

Torralba and Efros (2011) peformed an interesting experiment *‘Name that dataset’* and showed that simple model can correctly identify LiDAR images coming from same dataset. This is feasible as datasets appear to have strong intrinsic bias and thus, models trained on such data are able to learn a biased representations. They proposed fine-tuning strategies to compensate for such dataset biases without modifying the model training scheme.

Kou et al. (2021) developed a crowdsourcing data sampling framework for obtaining a ‘fair’ training dataset for face images. The sampling leverages an efficient batch-level demographic label inference model and a joint accuracy-aware data shuffling method. By removing less relevant images and increasing the demographic diversity, authors were able to train a state-ofthe-art model which achieve significant performance improvements. Models such as light CNN saw actual positive rate parity decrease from 0.282 to 0.055 when comparing a randomly sampled dataset to faircrowd while the accuracy and F1 scores remained comparable.

Clapes et al. (2018) identified that demographics (e.g. gender, race) introduced separate apparent biases for predicting person’s age using CNN model. They clustered individuals into known groups to obtain an apparent age estimate. By training models to predict apparent age followed by a constant adjustment term for each group, the authors reduced the 10% mean absolute error on their test dataset for real age prediction.

Howard et al. (2017) present a hierarchical approach that combines outputs from the Microsoft Emotion API algorithm with a specialized learner to reduce the bias of facial emotion recognition. They show that this methodology can increase overall recognition results by 17.3%. In the specialized learner, they used anthropometric features and also tune the model for ambiguous categories like fear and surprise.

#### 3.1.2. Representation Learning

Representation learning mainly focuses on automatically learning relevant representations of the data (typically as vectors/embeddings) by extracting useful information for building classifiers or other predictors Bengio et al. (2013). However, multiple previous works Morales et al. (2020); Zhang et al. (2021) have shown that advanced deep models could learn confounding features of non-task-related attributes and achieve good performance by overfitting. Such models are particularly bias towards the minority classes. This section discusses existing work where training scheme or model architecture is modified for ‘fair’ representation learning to reduce bias.

One popular theme is to develop ‘fair’ models for face images. Morales et al. (2020) proposed using privacy-preserving technique as a methodology to obtain fair models for face image classification (identity, attractiveness, and smiling). They developed SensitiveNets which forces the model to learn a ‘data representation’ with minimal demographic separation by using an adversarial regularizer and a series of linear blocks. The model’s effectiveness was compared against a model without regularization; overall accuracy decreased by 5% while adversarial tasks prediction accuracy (gender and ethnicity) dropped by 81% and 66%, respectively.

Alsulaimawi (2020) introduced a minimax adversarial framework, called features protector (FP), that aims to learn a new representation of the input data (face images) which maximize informativeness about the target features while being minimally informative to predict the sensitive features. Both criteria are satisfied only when the mutual information between input and target is maximized while minimizing mutual information between inputs and sensitive attribute. They observe a target AUC of 0.97 in the CelebA dataset while sensitive attribute Gender achieved close to random prediction (AUC 0.51).

Quadrianto et al. (2019) created an encoder/decoder architecture for enforcing fairness in ‘representation’ learning where the model will learn a mapping from an input domain to a fair target domain. Unlike other fair representation learning methods that learn latent embeddings, the method learns a representation that in the same space as the original input data. When comparing recognition ability in CelebA dataset overall accuracy decreased, yet achieved an increased TPR for the male demographic.

Zhang et al. (2021) investigated the development of a ‘fair’ camera-view agnostic model for person identification (re-ID) with minimal bias towards different camera perspectives. The ‘fair’ model was implemented using a view identification classifier branch preceded by a gradient reversal layer where the training with a reversal layer learn a ‘representation’ which is not influenced by the camera view. An ablation study that swapped DukeMTMC Ristani et al. (2016) and Market1501 Zheng et al. (2015) datasets for training and test sets revealed variable performance yet positive gains.

Similar to Zhang et al. (2021), Yu et al. (2018) developed an camera-view agnostic re-ID model but they designed an unsupervised asymmetric distance learning approach for cross-view clustering. The framework learnt the representation using cross-view cluster structure of Re-ID data, and thus help alleviate the view-specific bias. When evaluated on large re-ID datasets (e.g. VIPeR Gray et al. (2007), Market-1501 Zheng et al. (2015)), their model achieved higher performance than state-of-the-art models.

Jiang et al. (2019) explored the potential benefits of adding demographic attributes in an attribute-aware loss function to increase performance in facial recognition task. The attribute-aware loss function contains an additional term for an individual’s age and gender. Comparison of the model individual feature representation showed improved class separation amongst demographics resulting in improved identification quality across different gender and age groups.

To reduce the data distribution bias, Yan et al. (2019) developed a crossdomain aerial scene classification with limited target samples by using crossdomain distance metric learning. The authors advanced hybrid color features and bag of convolution features to force samples belonging to the same class to be more “similar” regardless of domain. They reported a decreases in class variability at inference and improvement in cross-domain performance.

Adeli et al. (2021) trained model with adversarial loss and showed a vanished correlation between the bias and the learned features on a synthetic dataset, a medical images (containing task bias), and a dataset for gender classification (containing dataset bias). The de-biased model resulted in representations that were invariant to the protected variable while obtaining comparable classification accuracy. For example on the medical dataset, their proposed approach resulted in the F1 score increasing from 0.68 to 0.74, while AUC only dropped from 80.8 to 80.9

Tonioni et al. (2019) developed a depth prediction model to generalize onto new domains by leveraging disparity measurements alongside with confidence estimators. They fine-tune both depth-from-stereo as well as depth-from-mono architectures by a novel confidence-guided loss function that handles the measured disparities as noisy labels weighted according to the estimated confidence. Weighting model losses across domains resulted in a fair model whose mean absolute error on the new domain decreased consistently.

Li et al. (2017) presented a low-rank parameterized CNN model for endto-end domain generalization learning. They outperformed new and older benchmark methods for a more complex mix domains of photo, sketch, cartoon and painting.

### 3.2 Clinical imaging study grouping

In comparison to natural images, slightly different debiasing trends for deep learning models have been observed for medical imaging – two key themes emerged from our review - (1) *multi-site* and (2) *demographic* fairness. Multi-site fairness focuses on model generalizability across different institutions/sites and remains a highly challenging issue for any model developer, especially for healthcare applications due to privacy policies. Lack of generalizability at the multi-institutional level is multi-faceted, involving various shifts in data such as demographics, imaging systems, and imaging acquisition and reconstruction protocols. Demographic fairness focuses on studying model performance for a single patient population with diverse racial/ethnic subgroups. Such studies are designed to observe the effects of demographic shifts at a refined level, understanding how patient demographics impact overall performance. By understanding demographic shifts, it is feasible to de-bias models at both - local and multi-institutional levels. The following sections will summarize latest developments for each theme.

#### 3.2.1. Multi-site fairness

In the existing literature, mostly optimal combinations of training datasets were employed for increasing variations in the training data to reduce institutional bias and achieve multi-site fairness.

Hägele et al. (n.d.) dealt with three types of common data related biases during model training - (1) biases which generates an allegedly good training performance whereas performances on independent datasets will be poor, (2) biases which are correlated with class labels and (3) sampling biases. They developed a model interpretation with pixel-wise heatmaps based on Layer-wise Relevance Propagation (LRP) Bach et al. (2015) technique assigns class relevance scores to individual pixels within the histopathology slides. Using the proposed visualization techniques, they visually evaluated potential dataset biases responsible for poor generalization. In parallel with uncovering these biases, authors eliminated the biased data points from the training data incrementally and showed that heatmaps offer an important tool to iteratively clean the dataset. Using modified cleaner training dataset, authors developed a model whose test performance increased by 0.05 AUC.

Cross- population test is a good way to measure of how robust an algorithm/model is. D. Das et al. (n.d.) utilized two chest X-ray datasets (with tuberculosis) from the Montgomery County, USA and Shenzhen, China to characterize model generalization on unseen populations. Test of the UStrained InceptionNet V3 model showed an overall AUC decrease from 0.84 to 0.79 on the dataset from China. Swapping US and Chaina test sets resulted in a smaller change in performance attributed to a larger dataset and a larger portion of positive findings. A separate model with same InceptionNet V3 architecture, trained on a mixture of the two datasets, achieved the highest overall performance. Again the improvements were attributed to having a greater prevalence of positive findings. The authors concluded that robust AI models can be built using larger cross-population datasets.

Similarly, Zech et al. (n.d.) studied models ability to detect pneumonia in chest radiographs by training and testing on data from different hospital systems in the US - National Institutes of Health Clinical Center (NIH; 112,120 from 30,805 patients), Mount Sinai Hospital (MSH; 42,396 from 12,904 patients), and Indiana University Network for Patient Care (IU; 3,807 from 3,683 patients). The prevalence of pneumonia was high enough at MSH (34.2%) relative to NIH and IU (1.2% and 1.0%) that merely sorting by hospital system achieved an AUC of 0.861 (95% CI 0.855–0.866) on the joint MSH–NIH dataset. Utilizing a training set with equal incidence across sites achieved the best performance on the testing set, suggesting balanced disease prevalence allows the model to learn disease specific features instead of site-related features.

Dinsdale et al. (n.d.) constructed a multi institutional AI model for detecting age on brain MR images. They identified that models could trivially assess the site of origin of the data with an accuracy of 96% and embeddings generated by the fully connected layer of the model also demonstrated clear separation based on scanner subtypes. They were able to improve classification performance by domain adaptation where they removed confounding factors by creating a feature space that is invariant to the acquisition scanner. In addition to the task specific loss, they used two additional losses to unlearn the domain features - (1) *domain loss* to assess how much domain information remains; (2) *confusion loss* that removes domain information by penalizing deviations from a uniform distribution. After this de-biasing, the models ability to identify site of origin decreased from 96% accuracy to just 56%. Meanwhile, on their task performance, age prediction error decreased from MAE of 4.10 to 3.90 in the OASIS dataset and 2.89 to 2.56 in the whitehall dataset. However, performance on the UK Biobank with de-biasing showed no improvements since the UK Biobank dataset has a much larger size for driving the initial performance of the network for normal training and with unlearning, all of the datasets finally able to influence the performance.

#### 3.2.2. Demographic fairness

One key unresolved question related to patient Health Insurance Portability and Accountability Act (HIPPA) is whether it is possible to re-identify patients based on their clinical information. Re-identification from data is not just a theoretical concept but has been demonstrated in several contexts, including clinical and social history datasets. For instance, demographics in an anonymized data set can function as a quasi-identifier that is capable of being used to re-identify individuals Sweeney (2002). The risk of patient re-identification and resultant privacy violation related legal ramifications can further increase the reluctance of allowing AI models access to comprehensive clinical datasets further hindering fair AI model development and evaluation.

Suriyakumar et al. (n.d.) sought to study the trade-offs between privacy-preserving techniques, such as differential privacy, and model utility and fairness. The authors demonstrate that differential privacy techniques (DP-SGD Abadi et al. (2016) - the de-facto approach for linear models and neural networks) guarantee of preserving model utility might not carry over to large medical datasets, such as predicting disease on chest x-rays. When applied to the NIH Chest X-Ray dataset, many positive disease labels were eliminated in the training process to preserve privacy. Despite operating on a large dataset, the removal of samples severely impacted model performance, reducing AUC from an average of 0.84 to 0.51. They also demonstrated that DP-SGD loses important information about minority classes (e.g., dying patients, minority ethnicities) that lie in the tails of the distribution. Having observed the impact on performance incurred by removing identifying information, it is of interest to explore the impact of quasi identifiers on the model performance, such as demographics.

Larrazabal et al. (2020) sought to study the impact of gender balance in medical imaging datasets used to train AI systems for computer-assisted diagnosis. They experimented with three deep neural network architectures and two well-known publicly available X-ray image datasets. Expectedly, the study shows that gender imbalance in chest x-ray datasets produces biased classifiers for diagnosis based on all models, with significantly lower performance in underrepresented groups. Interestingly, they did not find significant differences in performance between models trained with a gender-balanced dataset (50% male and 50% female) and an extremely imbalanced dataset from same gender. This suggests diversity of demographic attributes is important for improved model performance.

Shifts in training distribution may impact a model’s overall performance but the greatest burden of biased models are felt by protected subgroups for whom the model may fail to diagnose accurately. Seyyed-Kalantari et al. (n.d.) studied the impact of demographic biases in chest x-ray disease classification models using three large publicly available datasets. They quantified performance disparities by observing differences in true positive rates (TPR) among different protected demographic attributes such as patient sex, age, race, and insurance type which could be individually or in combination highly correlated with socioeconomic status. Authors justified the usage of TPR for measuring disparity as sick members of a protected subgroup with high TPR disparity would not be given correct diagnoses, even in an algorithm with high overall accuracy. For example, women were found to have significant differences in TPR despite contributing to 50% of the dataset. Further analysis of disease prevalence found varying levels of disparities in disease where women had an equal proportion of disease positive cases.

Guenther et al. (n.d.) investigated the impact of demographics on model performance while studying the classification of retinal fundus images for identification of age-related macular degeneration. The group used images from 68,400 individuals with available fundus images in the UK Biobank with additional manual classification in a subset of 2,013 participants, and found consistent miss-classifications related to physical characteristics. Upon further inspection, light eyes were found to have a 1.2% false positive rate whereas darker eye color and darker fundus images had a larger false positive rate of 3.0% when used for classification tasks.

## 4 Discussion

In this work, we performed a systematic review on ‘Fair’ AI models proposed for natural and medical images. Despite the large body of work focusing on AI model disparities in justice, law and education, there is little analytic work describing disparities in imaging applications. We screened 633 manuscripts, however only 22 relevant manuscripts were included in our analysis as mostly studies analyze the bias in the AI models without proposing any methodological solution. Among the 22 manuscripts, 14 manuscripts proposed solutions for natural images and focused two core themes - ‘fair’ training dataset (4/14) and representation learning (10/14). In debasing of medical image analysis, our review yielded 8 relevant manuscripts that carried two prevailing themes for medical images: model disparity across institutions (5/8) and model fairness with respect to patient demographics (3/8).

Debiasing techniques for natural images primarily focused on learning ‘less bias’ representation of the images for either classification of facial characteristics, age estimation, or developing a camera agnostic model for various image recognition tasks. Researchers have primarily explored three methodologies - (i) adversarial learning, (ii) joint learning and unlearning algorithm inspired in domain and task adaptation methods, and (iii) regularization of loss based on mutual information between feature embeddings and bias. Privacy-preserving approaches have also been proposed to disentangle certain attributes from learned representations. Clever sampling techniques and target task adjustment have also been proposed for reducing implicit bias in the training data.

While methods for natural images primarily focused on improving the three broad fairness criteria - demographic parity, equality of odds, and equality of opportunity - the primary aim of medical image debiasing is to improve cross-institutional fairness which involves understanding model generalization under large shifts in patient and imaging distributions. Not all institutions have the resources to train a state-of-the-art deep neural network for diseases prediction, and current business models treat algorithms as medical products that can be sold across health systems. Studies evaluating models on external datasets demonstrate consistent drops in performance. Inspection of these failures reveal models learn feature associations that are disease prevalence-dependent and sitespecific.

Even within a health system, there are several sources of variability (e.g. scanner type and patient demographics). From a clinician’s perspective, detection of disease should primarily focus on the pathology present in an image. Yet, several papers have demonstrated that models can learn spurious associations between features in classification and prediction tasks. Other works have attempted to hide demographic information from model predictions with dataset and/or model optimization, often producing inferior performance. Simpler approaches such as balancing training set across demographics improve cross-group performance. However, model performance on tests with equal proportions found bias across classes.

In an attempt to explore a relatively simpler way to improve model performance, several studies suggested that having a greater number of positive cases across demographics helped models perform better in validation. When evaluating the resulting performance, AUC is a good overall measure but does not accurately capture precision and recall within subpopulations. Newer metrics such as True Positive Rate (TPR) gap could provide a better indication of a model’s impact to patient care as it could be a surrogate for inherent model bias. Popular approaches to remove biases such as building demographic-specific models often suffer from a lack of demographic representation. The main challenge and goal of model debiasing is to reduce bias without the need for demographically balanced datasets. We observed that techniques that attempt to decouple demographic information and task predictions are not able to match baseline model performances and algorithmic development in medical imaging field is limited (Fig. **??**).

Because fairness in healthcare is a social problem, there is no simple solution or “silver bullet” that can “solve” fairness in AI systems. However, AI can play a role in developing fairer systems by reducing clinical decisions that are tainted by unconscious or conscious bias. Model development must reflect learned disease features and not patient demographic or imaging protocol to ensure robustness across diverse populations. Further exploration of demographic/institution agnostic models would be worthwhile to minimize disparity gaps. Once models are built, the second challenge is the reporting of performance metrics that estimate its impact on outcomes among marginalized groups in addition to accuracy. The use of the TPR gap and FPR comparisons may provide better insight into how the model may perform on different patient subgroups. AUC provides a good metric for overall model performance, but provides little intuition on how AI may widen the gap in outcomes across populations. More importantly, the current evaluation of fairness in AI models is still mainly focused on accuracy based on real-world data. If all we are aiming for is accuracy of predictions and classifications across populations, then the future world will be no different from today filled with inequities. If we want to use AI to prevent perpetuating or even magnifying health disparities, we need to come up with metrics that are not solely based on accuracy. How do we train and evaluate models when the ground truth is not fair? These are questions that will require the machine learning community to work more closely with social scientists. *Limitations*: Our systematic review is limited due to restriction to the imaging domain (and not general healthcare AI) and given our focus on papers that describe novel methodology for debiasing, we were limited to only examining a small set of eligible research studies. There exists a larger field of bias and fairness work that was excluded due to not being related to deep learning or fitting to our criteria. The topic of fairness itself is relatively new to AI applications, limiting our review to works published in the last five years. More research works may exist in the field through pre-prints that were excluded due to lack of peer review. Further analysis of biases was limited by papers only presenting overall performance metrics instead of group specific performance.

## Data Availability

The authors declare that all data supporting the findings of this study are available within the paper. Additional review data can be shared upon request in Covidence.

## 6 Authors Contribution

Concept and design: R.C., J.G., and I.B. Study selection: R.C., M.S and I.B. Data extraction: R.C., M.S and I.B. Drafting of the manuscript: R.C., M.S, J.G., B.P., and I.B. Critical revision of the manuscript for important intellectual content: H.T, B.P, L.C., Supervision: I.B.

